# Exploring of a novel monitoring tool for injury risk when sidestepping: Using unsupervised learning to characterize movement patterns

**DOI:** 10.1101/2022.11.06.22281978

**Authors:** Sina David, Gabor J. Barton

## Abstract

The monitoring of athletes is crucial to prevent injuries, identify fatigue or support return-to-play decisions. The purpose of this study was to explore the ability of Kohonen neural network self-organizing maps (SOM) to objectively visualize and characterize different movement patterns during sidestepping and to detect those patterns that are associated with signs of injury risk or fatigue. The marker trajectories of 631 pre-planned sidestepping trials were used to train a SOM. Out of 61913 input vectors, the SOM identified 1250 unique body postures, determined by the 3D marker positions. Visualizing the movement trajectories and adding the latent parameter time, allows for the investigation of different movement patterns. Additionally, the SOM can be used to identify zones with increased injury risk, by adding more latent parameters more directly linked to injuries which opens the option to monitor athletes and give feedback. The results highlight the ability of unsupervised learning to visualize movement patterns and to give further insight into an individual athlete’s status without the necessity to reduce the complexity of the data describing the movement.

## 1. Introduction

Active participation in sports is regarded as one contributor to life-long health including its social benefits ^1,2^. However, sport is also a major cause of injuries to the musculoskeletal system. These injuries result in loss of training hours and high socio-economic cost ^3^, may end an athlete’s career and in the worst-case result in degenerative diseases ^4^. Especially in more professional athletes, the narrow gap between performance enhancement and injury prevention makes it crucial to identify an athlete at risk. Various screening tools have been developed in the past years, but they often lack high reliability and validity or are not of high predictive value ^5^. In their meta-analysis, Bonazza et al. reported, that the Functional Movement Screen (FMS) which is a widely used screening tool for the prevention of musculoskeletal injuries, shows an intra- and inter-rater reliability of 0.81, which was rated as excellent. The predictive value of the tool showed that there was a 2.74 times higher chance of getting injured if scoring lower than 14 in the FMS. However, concerns remained concerning validity ^6^. Also, the question remains how sensitive the tool is to detect an actual athlete at risk ^5^.

Recent studies showed the impact of individual movement patterns on injury-related structural loadings during fast sidestepping manoeuvres ^7–9^. The results provided a clear indication that preparatory strategies such as trunk inclination, preorientation and foot strike pattern determine the knee valgus moment which is a proxy for anterior cruciate ligament (ACL) load. Also, the effects of fatigue on the kinematics and kinetics during fast sidestepping were investigated. Several studies reported changes in hip and knee flexion angles ^10–12^, an increase of the knee valgus angle ^10,12^ as well as changes in the ground reaction forces ^11^. These suspected risk factors, with reasonably good sensitivity and specificity ^13,14^, were often used to inform screening models that were hoped to identify athletes at risk. However, most of these models require the definition of a threshold for the used predictor variables to detect athletes at risk. This is in contrast to the idea that the transition between no risk and risk is not categorical but continuous and that athletes who score similarly should not end up in different groups if they happen to fall on the two sides of the division line.

However, despite such advances in knowledge, successful prevention could not be achieved ^5^. The nature of injuries is complex and is influenced by internal and external risk factors in addition to the inciting event with all its factors ^15,16^. Modern lab technology allows researchers to investigate such complex problems from different perspectives. Three-dimensional movement analysis systems can capture accurate positions and orientations of the body and the use of inverse dynamics can combine the movement data with external forces to calculate joint loading. Additional features like electromyography to obtain muscle activity, pressure distribution within the shoe and others can be added to further increase the amount of data and to give a more granular picture. This results in a large amount of multidimensional data which is a big advantage of modern data acquisition, but the problem arises of how to process and more importantly understand and connect all the data. The human brain can receive, process, and remember between seven to nine items at a time ^17^. When reviewing the large datasets gathered through motion analysis, only the lower limb joint angles of the right leg in three dimensions already exceed this number of items. So, the researcher must decide on a preselection of parameters. From other subdisciplines employing 3D motion analysis, it was reported that these assumptions are dependent on the profession or experience of the investigator or even the institution the researcher works at ^18,19^. In sports biomechanics, the selection is often based on the mechanical understanding of the load-tissue interaction. However, every movement is the result of the interplay of mechanics, tissue characteristics, motor control, psychology, and others. It might therefore be impossible to identify a single cause that forces an athlete into a movement pattern with an elevated risk of injury while others will end up using a different pattern in the same situation. Additionally, the transition from one movement pattern into another is continuous - as are the risk factors, which makes data analysis even more complex and the grouping of athletes to further investigate them is neglecting athlete-specific factors ^5^.

To summarise, the state-of-the-art motion analysis approaches result in high-quality datasets. The conventional ways to approach these datasets require a reduction of the amount of data and often also force the researcher to make a-priori assumptions. This may result in a narrowed view of the dataset and eventually lead to overestimating the importance of single parameters together with loss of essential information. One solution to the raised issues is neural networks through Kohonen self-organizing maps ^20^. These unsupervised neural networks can overcome the described problems as they can process large quantities of input data without the need to make an a-priory reduction of data ^21^.

The aim of this study was therefore to explore the ability of SOMs to visualize movement patterns during sidestepping. Further, it was hypothesized, that the SOM can identify typical movement patterns during sidestepping, some of which are associated with injuries.

## 2. Material and Methods

Lower body marker trajectories of athletes performing planned full-effort 90° sidestepping ^22^ were used to train an unsupervised neural network using Kohonen self-organizing maps ^20^.

### 2.1 Participants

The dataset contained 613 trials of 67 athletes in total (adults: 26 male, 31 female, age 22.6 ± 3.3 years, height 1.77 ± 0.1 m, mass 70.9 ± 0.1 kg; children: 10, age 9.8 ± 1.0 years, height 1.45 ± 0.1 m, mass 36.98 ± 6.38 kg). All participants were free of injury or pain and gave their written informed consent to participate in the study. Ethical approval for the study was given by the University’s ethical committee.

### 2.2 Data collection

The trajectories of 28 lower body retro-reflective markers were captured by 14 infrared cameras (sampled at 200 Hz, VICON, Oxford, UK). Two floor-embedded force plates (Kistler, Winterthur) recorded the ground reaction forces (GRF) of the execution (EXEC) and depart (DEPART) contact phases of the sidestepping task. Each athlete performed sidesteps using their preferred and non-preferred leg as the EXEC leg, where the preferred side was determined by asking them to kick a ball.

Inverse kinematics and dynamics were carried out using AnyBody (V6, Aalborg, Denmark) to calculate the 3D lower-limb joint angles and internal joint moments. For this purpose, kinematic and kinetic data were filtered with a recursive 2nd order low pass filter with a cut-off frequency of 20 Hz^23^.

### 2.3 Data postprocessing

MATLAB 2020b (The Mathworks) was used for all the following steps. As the dataset contained sidestepping to the left and the right, the marker positions were mirrored, and the GRF and GRF moments were rotated around the global reference frame which was placed centrally between the two force plates in case athletes turned to the left and used force plate 2 for the EXEC contact. After this procedure, all sidesteps were executed to the right, with the left leg touching Force Plate No.1 during the EXEC contact and the right leg touching Force Plate No. 2 during the DEPART contact. The global reference frame was placed between the two force plates (see Figure 1). This process ensured, that the network was not biased by sidestepping either to the left or the right side.

**Figure 1:**
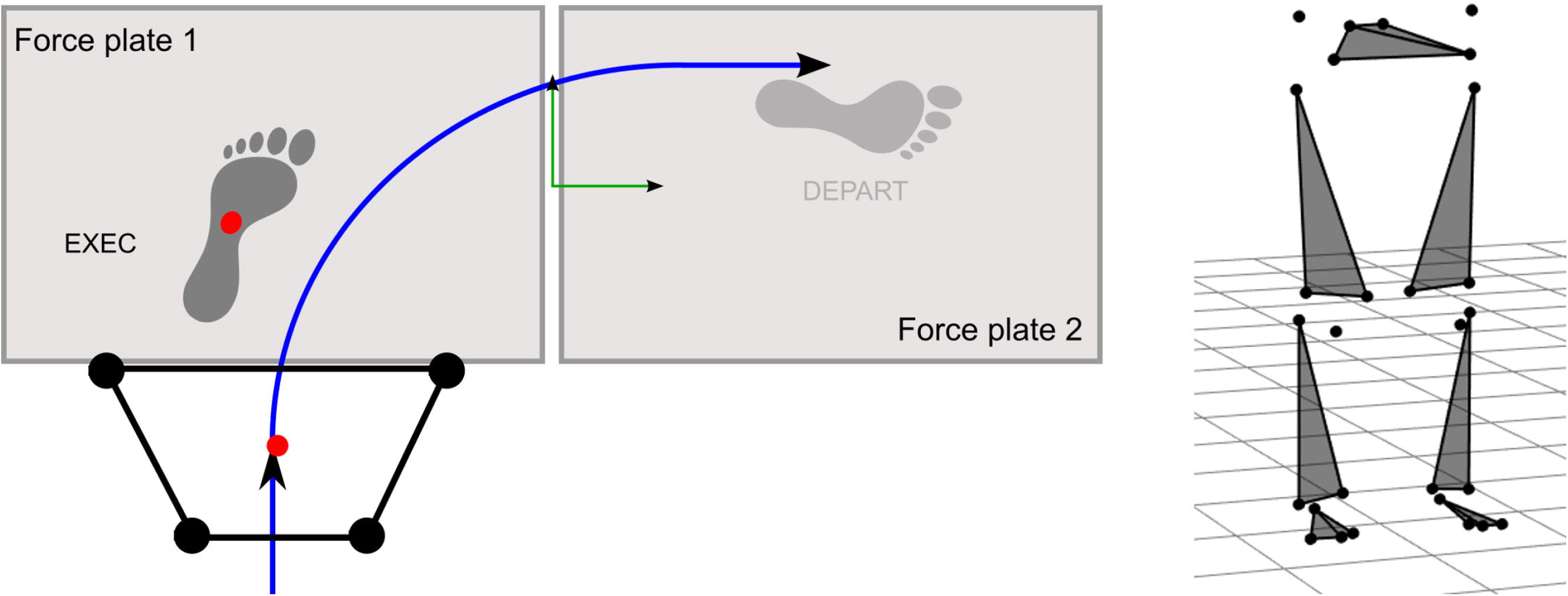
Left: Laboratory setup. All sidestepping trials that were executed to the left side were modified to match the right sidestepping trials by means of using the left leg as the EXEC leg and touching force plate 1 during the EXEC contact. The black trapezium represents the pelvis segment with the four pelvis markers that were used to calculate the pelvis center at initial touch-down of the EXEC contact. The green coordinate system represents the global reference frame. Right: 3D position of the lower-body markers on the pelvis, thighs, shanks and feet.

The marker trajectories, joint angles and moments and the GRF were time-normalized to the ground contact of the EXEC and DEPART contact, using the vertical GRF. Touch down and take off were the first and last instances where the GRF crossed a threshold of 20 N.

To test the SOM’s ability to be used as a tool to screen the ACL-relevant load that is related to specific movement patterns, we extracted ACL injury-relevant factors by means of knee abduction moment, and knee and hip joint angles to account for the leg alignment and used them as latent variables after having trained a SOM with the lower-body marker trajectories of athletes during sidestepping. The idea behind this is to identify movement patterns, that will cross areas of increased injury risk of ACL-relevant joint loading.

In the next step, all lower-limb marker trajectories were space-corrected. To ensure that the distance from the global reference frame does not bias the input data for the neural network, the centre of the four pelvis markers at the instance of the touch-down of the EXEC contact was calculated and subtracted from the time-normalized marker trajectories (**Error! Reference source not found**.Figure 1). With this, the confounding effect of the absolute position of the body in the global reference frame was removed.

Finally, one matrix containing all 631 trials and the 84 marker trajectories (61913 × 84, 613 trials x 101 data points (613×101=61913)) was concatenated to generate the input data sets for the neural network.

### 2.4 Network Specifications and Workflow

An unsupervised learning Kohonen self-organizing map (SOM) neural network was trained with the marker trajectories (Kohonen, 2001) using the SOM Matlab Toolbox (V 2.1). The input layer consisted of a matrix of 61913 input vectors. The initial connection weights of the neurons are set by the first two principal components of the input dataset. The following section describes the training process of the SOM:

(1) Normalization of the input vectors to zero mean and unit variance and defining the number of nodes on the SOM. (2) To train the SOM, the algorithm iteratively chooses one random input vector X and calculates the Euclidean distance ε to all neurons of the Kohonen layer, where each of the neurons is represented by an n-dimensional weight vector W, with n being equal to the number of input vectors (**Error! Reference source not found**.). The neuron with the smallest Euclidean distance ε is called the winner neuron. (3) The weight W of the winner neuron r is adjusted by using the following equation:

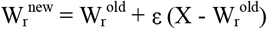

Geometrically this means that the weight vector W of the winner neuron is moving one step of the length of ε closer towards the chosen input vector. A Gaussian neighbourhood relationship ensures that the weights of neighbouring neurons are also reduced. (4) The learning procedure is stopped if no further significant changes in the weight vectors are detected.

## 3. Results

After initialization, the Kohonen layer consisted of 1247 neurons (determined by the size of the input matrix), distributed over a 43 × 29 rectangular map with a hexagonal neighbourhood topology of each neuron (Figure 2). This means that the 61913 input postures were reduced to 1247 relevant movement patterns to describe the whole data set. From the 2D trajectory of the best-matching neurons on the map (see Figure 2) one can reconstruct the movement of a specific input trial from the codebook vectors representing the movement patterns in the SOM weights. With this, the movement trajectory of an athlete can be followed and also compared to either other athletes, or within the same athlete to monitor changes. From Figure 3B it becomes visible that the movement trajectory of four different athletes follows quite distinct paths on the 2D SOM.

**Figure 2:**
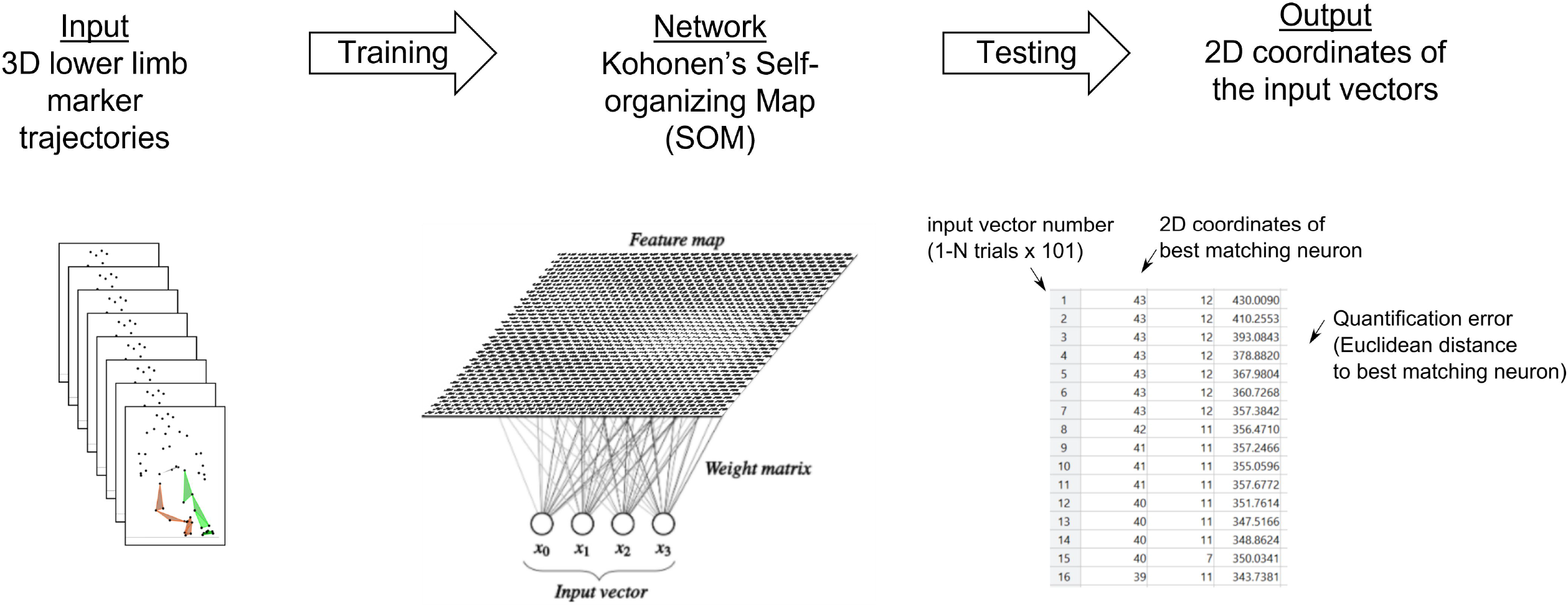
Workflow

**Figure 3:**
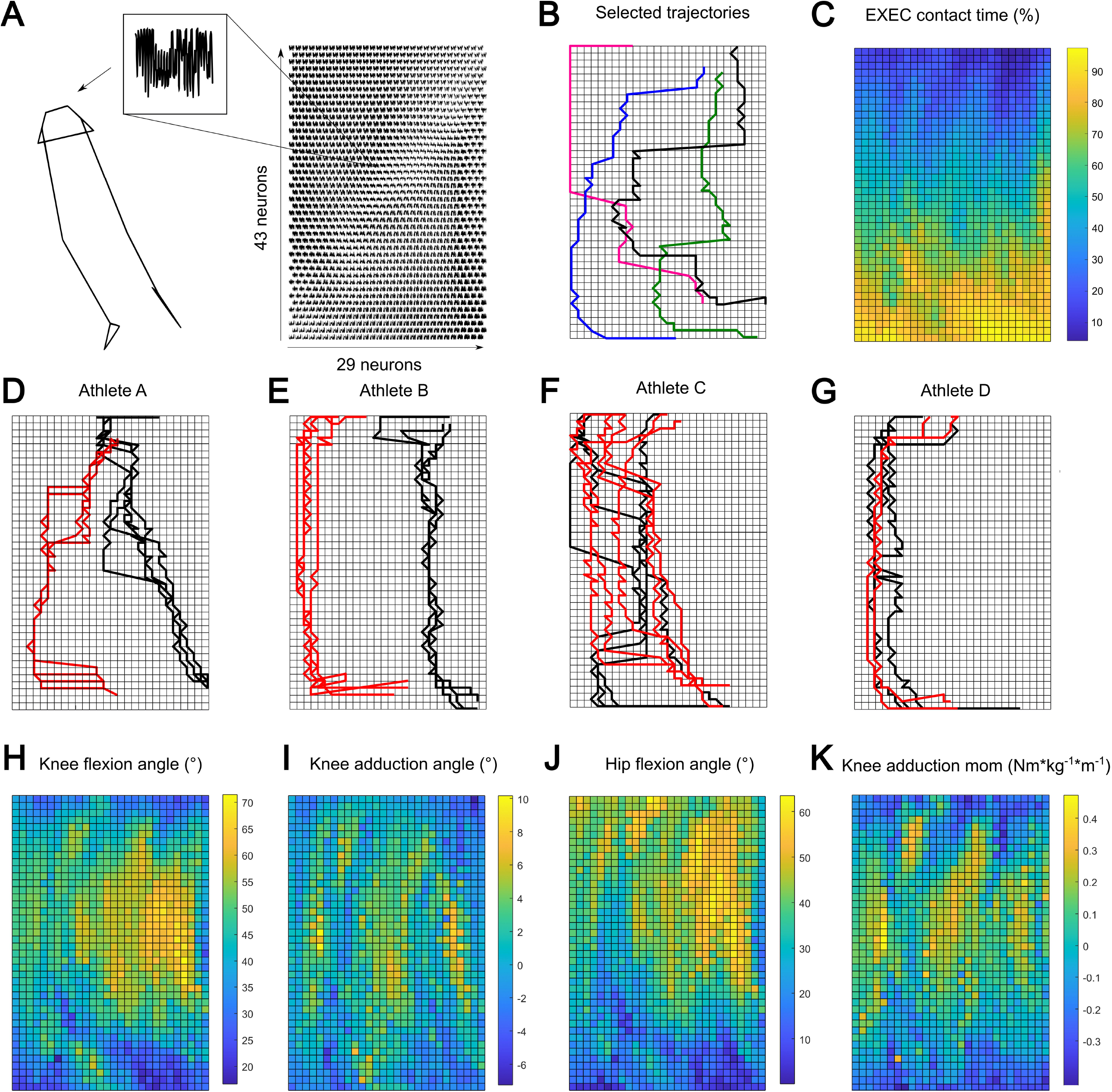
Results. A: plane plot of the SOM showing the content of the single neurons that can be reconstructed into a stick figure B: movement trajectories of four selected athletes, showing distinct paths on the SOM. C: Adding the timing during the EXEC contact (%) as a latent feature to the SOM. D-G: Visualization of the movement trajectories of sidestepping trials towards the preferred (black) and non-preferred (red) side of four randomly selected athletes. H-K: Adding the knee flexion and adduction angle, hip flexion angle and knee adduction moment as latent features to the SOM.

In combination with the 2D coordinates and the quantification error (see Figure 2), additional latent features can be used to label the SOM. The first latent feature was the time instance (%) during the EXEC contact. As can be deducted from Figure 3C, the distribution of the postures over the SOM is very smooth with regard to their occurrence in time during the EXEC contact. With this, zones on the map that are interesting e.g. in terms of injury risk can be identified. Most non-contact injuries occur during the weight acceptance phase which refers to approximately the first 30% of the contact phase (the upper part of the SOM in Figure 3 C). Also, the number of neurons associated with each of the phases during the ground contact is different between athletes, as can be appreciated from Figure 3B. While the pink trajectory shows the highest resolution (number of neurons per phase) during the first 40 % of the contact, the black trajectory is showing a higher resolution during 50 to 60 % and the green one dominates the last 20% of the contact phase. This directly translates into the number of distinct postures that were covered per phase. Taking this one step further, changes to the movement trajectory within one athlete can be monitored. The maps in Figure 3D-G show the trajectories of four different athletes and the trials towards their preferred (black) and non-preferred (red) direction. While athlete A and B show clear changes to their movement trajectories, athletes C and D do not seem to be affected by changing the movement direction. The same procedure can be done with data pre- and post-injury, or to monitor fatigue.

Finally, adding more latent features such as joint angles or moments can be used to interpret the path of the trajectory. Maps in Figure 3H-K display the distribution of the knee’s sagittal and frontal and the hip’s sagittal joint angle as well as the frontal knee joint moment. These features were chosen for illustration as they are related to knee joint injuries. By projecting the trajectories on these maps, it becomes possible to investigate, whether an athlete crosses an area of high joint loading or joint positions with a high risk of injury. Also, the aforementioned changes in the path of the trajectory can directly be translated into changes in the joint angles and moments. The change of movement path of athletes A and B from the preferred to the non-preferred side results in a decrease in knee and hip flexion and an increase in knee abduction, which is more likely to result in a collapse of the knee joint. Athletes C and D show this movement strategy for all their trials which could indicate that they are in general at higher risk for knee joint injuries.

## 4. Discussion

The results of this study demonstrated the power of the SOM to reduce the dimensionality of a large dataset of 61913 input vectors to 1247. This is done with some loss of detail while retaining most of the information content as proven by the ability to reconstruct an entire movement pattern from the stored vectors of the SOM. Further, this is a data-driven approach, which did not require any assumptions or hypotheses for data reduction.

The visualization of the marker trajectories during sidestepping using a Kohonen self-organising map allows for a new way to investigate individual movement patterns and the associated research questions. The SOM itself can be used to understand the distribution of the different postures along the individual movement trajectories. The option to add other latent parameters such as time gives insight into the proportion of time taken within the different phases of the movement. This does not mean that an athlete spends more time in the weight acceptance phase, but the resolution of the SOM for this athlete is higher in this phase (indicated by the number of neurons in this phase), meaning that the differences between the single input vectors are higher for this athlete than for the other examples chosen for Figure 3B. By following the black trajectory in Figure 3A, it can be observed that the athlete shows more knee and hip flexion of the execution leg than the other athletes, suggesting that a higher number of different postures were achieved. Adding the lower limb joint angles as latent variables as done with time, could be one way of getting deeper insight into these individual movement patterns. It has to be mentioned that time or joint angles were not included in the input vectors, thus the smooth distribution of the timing (Figure 3C) confirms the quality of the learning process.

Taking this one step further, labels that are associated with injury risk such as joint moments can also be added as latent variables. With this, it becomes possible to identify zones on the map, where several risk factors fall together, such as a high knee valgus moment during the weight acceptance phase ^8,9,24^. A movement trajectory that is travelling through this zone could identify an athlete with a higher risk of injury. This approach is completely different from the screening tools that are commonly used. There is no need to define thresholds for joint loading or other risk factors, which was critically discussed recently ^5^. An athlete that is closer to the centre of a risk zone will be more likely to be exposed to an injury-relevant load. Also, there is no need to group athletes according to any hypotheses, which always contain the risk of missing effects or choosing a grouping variable that is not discriminative. Therefore, SOMs are a sensitive tool, to highlight individual movement patterns and given their mode of operation offer the identification of risk.

A distinct advantage of the proposed method is the gradual change that is distributed across the map. With this, the effect of small changes in the movement on the overall movement pattern can be visualized. Another big benefit of the proposed method is its flexibility. If the research question focuses on the ankle instead of knee injuries, the same map can be used but labelled with ankle-relevant latent parameters. Also, the SOM can be used to monitor athletes, by observing how their trajectories on the 2D SOM change over time, e.g., indicating a movement towards or away from identified risk zones. The advances in pose estimation would allow feeding the network with other features such as joint angles or even parameterised images. For example, this could be used for direct feedback to the athlete in form of warnings if risky postures are detected repeatedly.

It has to be mentioned that the used data set does not contain data from injured athletes. Therefore, no conclusion can be drawn towards the predictive value of the SOM as an injury screening tool. However, this was also not the aim of the study. Also, we only included the lower-body marker trajectories as upper-body markers were not present for all included athletes. Where whole-body movements contribute to injury risk, the additional data could improve the sensitivity of the method.

## 5. Conclusion

Kohonen’s self-organizing maps are a useful method to investigate the outcome of different movement patterns without the need for assumptions or grouping of athletes according to global parameters. They can highlight how small adaptations in the movement pattern influence the movement pattern. In combination with the advances in posture estimation, they can be used for feedback training or athlete evaluation.

## Data Availability

All data produced in the present study are available upon reasonable request to the authors

## References

1. Biddle S, Fox KR, Boutcher SH. Physical Activity and Psychological Well-Being. Vol 552. Routledge London; 2000.

2. Hagger M, Chatzisarantis N. The Social Psychology of Exercise and Sport. McGraw-Hill Education (UK); 2005.

3. Gottlob CA, Baker CL, Pellissier JM, Colvin L. Cost effectiveness of anterior cruciate ligament reconstruction in young adults. Clin Orthop Relat Res. 1999;(367):272–282.

4. Ruiz AL, Kelly M, Nutton RW. Arthroscopic ACL reconstruction: a 5–9 year follow-up. Knee. 2002;9(3):197–200. doi:10.1016/S0968-0160(02)00019-4

5. Bahr R. Why screening tests to predict injury do not work-and probably never will…: a critical review. Br J Sports Med. 2016;50(13):776–780. doi:10.1136/bjsports-2016-096256

6. Bonazza NA, Smuin D, Onks CA, Silvis ML, Dhawan A. Reliability, Validity, and Injury Predictive Value of the Functional Movement Screen: A Systematic Review and Meta-analysis. Am J Sports Med. 2017;45(3):725–732. doi:10.1177/0363546516641937

7. David S, Mundt M, Komnik I, Potthast W. Understanding cutting maneuvers – The mechanical consequence of preparatory strategies and foot strike pattern. Hum Mov Sci. 2018;62:202–210. doi:10.1016/j.humov.2018.10.005

8. Dempsey AR, Lloyd DG, Elliott BC, Steele JR, Munro BJ. Changing sidestep cutting technique reduces knee valgus loading. Am J Sports Med. 2009;37(11):2194–2200. doi:10.1177/0363546509334373

9. Donnelly CJ, Chinnasee C, Weir G, Sasimontonkul S, Alderson J. Joint dynamics of rear- and fore-foot unplanned sidestepping. J Sci Med Sport. 2017;20(1):32–37. doi:10.1016/j.jsams.2016.06.002

10. Zago M, David S, Bertozzi F, et al. Fatigue Induced by Repeated Changes of Direction in Élite Female Football (Soccer) Players: Impact on Lower Limb Biomechanics and Implications for ACL Injury Prevention. Front Bioeng Biotechnol. 2021;9. doi:10.3389/fbioe.2021.666841

11. Iguchi J, Tateuchi H, Taniguchi M, Ichihashi N. The effect of sex and fatigue on lower limb kinematics, kinetics, and muscle activity during unanticipated side-step cutting. Knee Surgery, Sports Traumatology, Arthroscopy. 2014;22(1):41–48. doi:10.1007/s00167-013-2526-8

12. Hosseini E, Daneshjoo A, Sahebozamani M, Behm D. The effects of fatigue on knee kinematics during unanticipated change of direction in adolescent girl athletes: a comparison between dominant and non-dominant legs. Sports Biomech. Published online June 14, 2021:1–10. doi:10.1080/14763141.2021.1925732

13. Myer GD, Brent JL, Ford KR, Hewett TE. Real-Time Assessment and Neuromuscular Training Feedback Techniques to Prevent Anterior Cruciate Ligament Injury in Female Athletes. Strength Cond J. 2011;33(3):21–35. doi:10.1519/SSC.0b013e318213afa8

14. Hewett TE, Myer GD, Ford KR, et al. Biomechanical measures of neuromuscular control and valgus loading of the knee predict anterior cruciate ligament injury risk in female athletes: a prospective study. Am J Sports Med. 2005;33:492–501. doi:10.1177/0363546504269591

15. van Mechelen W, Hlobil H, Kemper HCG. Incidence, Severity, Aetiology and Prevention of Sports Injuries. Sports Medicine. 1992;14(2). doi:10.2165/00007256-199214020-00002

16. Bahr R, Krosshaug T. Understanding injury mechanisms: a key component of preventing injuries in sport. Br J Sports Med. 2005;39(6):324–329. doi:10.1136/bjsm.2005.018341

17. Miller GA. The magical number seven, plus or minus two: Some limits on our capacity for processing information. Psychol Rev. 1956;63(2). doi:10.1037/h0043158

18. Skaggs DL, Rethlefsen SA, Kay RM, Dennis SW, Reynolds RAK, Tolo VT. Variability in gait analysis interpretation. Journal of Pediatric Orthopaedics. 2000;20(6):759–764.

19. Watts HG. Gait laboratory analysis for preoperative decision making in spastic cerebral palsy: is it all it’s cracked up to be? J Pediatr Orthop. 1994;14(6):703–704.

20. Kohonen T. Self-Organizing Maps. Vol 30. Springer Berlin Heidelberg; 2001. doi:10.1007/978-3-642-56927-2

21. Barton G, Lees A, Lisboa P, Attfield S. Visualisation of gait data with Kohonen self-organising neural maps. Gait Posture. 2006;24(1). doi:10.1016/j.gaitpost.2005.07.005

22. David S, Komnik I, Peters M, Funken J, Potthast W. Identification and risk estimation of movement strategies during cutting maneuvers. J Sci Med Sport. 2017;20(12):1075–1080. doi:10.1016/j.jsams.2017.05.011

23. Kristianslund E, Krosshaug T, van den Bogert AJ. Effect of low pass filtering on joint moments from inverse dynamics: implications for injury prevention. J Biomech. 2012;45(4):666–671. doi:10.1016/j.jbiomech.2011.12.011

24. Boden BP, Sheehan FT, Torg JS, Hewett TE. Non-contact ACL injuries: mechanisms and risk factors. J Am Acad Orthop Surg. 2010;18(9):520.

